# Transfer Learning for Medical Imaging: An Empirical Evaluation of CNN Architectures on Chest Radiographs

**DOI:** 10.64898/2026.01.07.26343591

**Authors:** Harsh Santosh Salve

## Abstract

This paper presents a comprehensive comparative study of five state-of-the-art CNN architectures, VGG19, ResNet50, InceptionV3, DenseNet121, and EfficientNetB0 for multi-class classification of Chest X-ray images (CXR) into four categories: Edema, Normal, Pneumonia, and Tuberculosis (TB). The models were trained, validated, and tested on a dataset comprising 6,092 training and 325 testing images across four distinct classes. Each architecture was initialized with ImageNet weights, augmented with a custom classifier, and fine-tuned under identical conditions to ensure a fair comparison. The models are evaluated on a comprehensive set of metrics, including accuracy, per-class recall, training time, and model complexity. Experimental results indicate that VGG19 achieved the highest classification accuracy of 98.15%, followed closely by ResNet50 at 97.54%. This study provides empirical evidence to guide the selection of appropriate deep learning models for chest X-ray diagnosis, balancing performance with operational constraints

## I. Introduction

The integration of artificial intelligence in healthcare, especially in radiology for early and accurate diagnosis, is crucial for effective treatment and transforming the diagnostic process. Chest X-rays are the most common and cost-effective medical imaging modality for screening and diagnosing. However, the manual interpretation of CXRs is time-consuming and subject to inter-observer variability. This process can be subjective, time-consuming, and prone to error. Automated classification systems based on deep learning offer a promising solution to augment radiologists capabilities, improve diagnostic speed, and enhance accuracy.

Convolution Neural Network(CNN) has become the cornerstone of modern computer vision task, CNN can automatically learn hierarchical features from images, often surpassing human-level performance in specific visual recognition tasks. Architectures like VGG, ResNet, Inception, DenseNet, and EfficientNet have demonstrated exceptional performance on large-scale datasets. Leveraging these pre-trained models via transfer learning is a widely adopted strategy in medical imaging, where labelled data is often scarce in the medical domain. Transfer learning provides an effective solution to this problem. As numerous studies have applied individual models to CXR classification, a systematic comparison under a unified experimental setup is necessary to assess their strengths and weaknesses objectively

This paper aims to fill this gap by conducting a rigorous empirical comparison of five prominent CNN architectures for multi-class CXR classification. The primary contributions of this work are:

1. Implementing a standardized pipeline to train and evaluate VGG19, ResNet50, InceptionV3, DenseNet121, and EfficientNetB0 on the same CXR dataset
2. multi-faceted performance analysis encompassing overall accuracy, training efficiency, model complexity, and class-wise performance
3. Providing insights into the performance-accuracy trade-offs to aid researchers and practitioners in selecting the most suitable model for their specific clinical application.

## II. RELATED WORK

### A. Deep Learning for Chest X-ray Analysis (Heading 2)

Deep learning for chest X-ray analysis has advanced significantly with the release of large-scale datasets like NIH ChestX-ray8 [1] and CheXpert [2]. Seminal work by Rajpurkar et al. [3] demonstrated radiologist-level pneumonia detection, establishing transfer learning as a standard practice. While recent methods incorporate interpretability tools like Grad-CAM [4], systematic comparisons of multiple architectures for multi-class pulmonary disease classification remain limited.

### B. Individual CNN Architecture Evaluations

The evaluated architectures represent key advancements in CNN design: VGG19 [5] established the efficacy of deep convolutional stacks; ResNet [6] enabled the training of very deep networks via skip connections; InceptionV3 [7] optimized computational efficiency through factorized convolutions; DenseNet [8] promoted feature reuse through dense connectivity; and EfficientNet [9] automated architecture scaling for optimal performance. Each offers distinct advantages for medical imaging, depending on dataset characteristics and computational constraints.

### C. Comparative Studies in Medical Imaging

Comparative CNN evaluations in medical imaging remain limited, with studies often focusing on binary classification [10] or multi-label scenarios [11]. Methodological inconsistencies further hinder fair comparisons. This study addresses these gaps by systematically comparing five foundational architectures under standardized conditions for a four-class pulmonary disease classification task, providing empirical guidance for clinical deployment.

## III. Methodology

This section details the dataset, preprocessing steps, and the experimental setup used to train and evaluate the models.

### A. Dataset and Preprocessing

The study utilized a dataset containing 6,417 chest X-ray images, split into 6,092 for training and 325 for testing (validation) images. These images were categorized into four distinct diagnostic classes: Edema, Normal, Pneumonia, and Tuberculosis. The distribution of data across these classes provides a foundation for multi-class classification. To ensure compatibility with the pre-trained model, all images were resized to the required input dimensions of each network (224×224 for most, 299×299 for InceptionV3). Data augmentation techniques, including random shearing, zooming, and horizontal flipping, were applied to the training set to increase variability and reduce overfitting. This was achieved by applying data augmentation to the training set using the ImageDataGenerator class from Keras. The augmentations included Shear Rage: 0.2, Zoom Range: 0.2, and Horizontal Flip: True.

A critical preprocessing step was the inclusion of model-specific normalization. Each model’s dedicated preprocessing function (e.g., vgg19.preprocess_input) was integrated directly into the model architecture using a Lambda layer. This ensures that the input images are normalized according to the same statistics used when the models were originally trained on ImageNet.

### B. Model Architectures

1. VGG19: The VGG19 architecture is defined by its structural homogeneity and significant depth for its time, comprising 16 convolutional and 3 fully-connected layers. Its core design principle is the systematic use of small 3×3 convolutional filters with a stride and padding of 1, arranged in sequential stacks. This small receptive field size increases the network’s depth and non-linearity (via the ReLU activation function, f(x) = max(0, x)) while maintaining the same effective receptive field as a larger single filter, but with fewer parameters. Spatial down-sampling is achieved exclusively through 2×2 max-pooling layers with a stride of The network culminates in two fully-connected (FC) layers with 4,096 units each, followed by a softmax output layer. A key regularization technique employed is Dropout, which randomly sets a portion of activations to zero during training to prevent overfitting by reducing complex co-adaptations between neurons. While highly effective in feature extraction, its reliance on large FC layers results in a high computational cost and a substantial number of parameters (over 20 million), making it relatively inefficient compared to later architectures
2. ResNet50: The Residual Network (ResNet) architecture introduced a paradigm shift to enable the training of very deep networks by addressing the vanishing gradient problem. Its fundamental innovation is the residual block and the use of skip connections (or identity mappings). Instead of a stack of layers learning a desired underlying mapping H(x), they are designed to learn a residual function F(x):= H(x) - x. The original input x is then added to the output of the block via a shortcut, yielding H(x) = F(x) + x. This element-wise addition allows gradients to flow directly through the network via these identity paths, effectively bypassing the non-linear transformation layers during backpropagation. This prevents the gradient from becoming indefinitely small and enables stable training of networks with hundreds of layers. The ResNet50 variant specifically employs a “bottleneck” design within its residual blocks, which uses a sequence of 1×1, 3×3, and 1×1 convolutions. The initial 1×1 convolution reduces dimensionality (channels), making the 3×3 convolution less computationally expensive, and the final 1×1 convolution restores dimensionality. This efficient design allows ResNet50 to achieve superior performance with 50 layers.
3. InceptionV3: As an evolution of the Inception family, InceptionV3 was designed to maximize computational efficiency and representational power within strict resource constraints. Its core innovation lies in the aggressive use of factorized convolutions. This involves replacing large filters with smaller, more efficient asymmetric convolutions. For instance, a 5×5 convolution is decomposed into two consecutive 3×3 convolutions, reducing parameters by 28%. Further, a 3×3 convolution can be factorized into a 1×3 followed by a 3×1 convolution. This asymmetric spatial factorization significantly reduces computational cost while expanding the network’s width and feature diversity within each module. Additionally, InceptionV3 incorporates auxiliary classifiers attached to intermediate layers. These classifiers propagate the gradient directly to the earlier layers during training, acting as a regularizer and further combating the vanishing gradient problem. The model also utilizes techniques like label smoothing, a regularization loss function that discourages the model from making overconfident predictions. With 48 layers, it achieves a powerful balance between depth, width, and efficient resource utilization.
4. DenseNet121: DenseNet (Dense Convolutional Network) radically rethinks connectivity by introducing dense blocks. Within each block, every layer receives the feature maps from all preceding layers as input and passes its own feature maps to all subsequent layers. This is achieved via depth-wise concatenation. The l-th layer has l inputs, consisting of the feature maps from all previous layers, and its own feature maps are passed on to (L-l) subsequent layers. This architecture promotes several key advantages: it encourages feature reuse, drastically reduces the number of parameters (as each layer can be very narrow, producing only a small set of feature maps, known as the growth rate), and strengthens gradient flow throughout the network. Transition layers between dense blocks, typically consisting of a 1×1 convolution (for feature compression) and a 2×2 average pooling layer, control the growth of feature map dimensions. This dense connectivity pattern ensures that information from the initial layers can flow directly to deeper layers, making the network highly parameter-efficient and mitigating information loss.
5. EfficientNetB0: EfficientNet represents a systematic approach to model scaling. Previous models scaled dimensions like depth (number of layers), width (number of channels), or resolution (input image size) arbitrarily. EfficientNet, however, introduces a compound scaling method that uses a fixed set of scaling coefficients to uniformly scale all three dimensions, depth, width, and resolution, in a balanced way. The rationale is that these dimensions are interdependent; increasing resolution, for instance, should be accompanied by a corresponding increase in network depth and width to effectively capture finer-grained patterns. The baseline architecture, EfficientNet-B0, was itself discovered through Neural Architecture Search (NAS) using the AutoML MNAS framework, optimized for both accuracy and floating-point operations (FLOPS). The resulting model employs mobile inverted bottleneck convolution (MBConv) blocks, similar to those in MobileNetV2, which utilize depthwise separable convolutions and squeeze-and-excitation blocks for channel-wise attention. This compound scaling enables EfficientNet-B0 to achieve state-of-the-art performance with an order of magnitude fewer parameters and FLOPS than previous models, making it exceptionally efficient for both training and inference.

## IV. Training Strategy and Hyperparameters

This section outlines the training methodology, model compilation, and hyperparameter selection used in the comparative study.

### A. Transfer Learning Approach

A **transfer learning** methodology was adopted for all five CNN architectures. This approach is standard practice in medical imaging, where datasets are often too small to train deep, complex networks from scratch without significant overfitting

1. ***Justification***: *By using models pre-trained on the large-scale ImageNet dataset, we leverage a rich hierarchy of learned visual features (e*.*g*., *edges, textures, shapes). These low-level features are highly transferable to the domain of medical imaging*.
2. ***Implementation:*** *For each model, the base convolutional layers were loaded with their pre-trained ImageNet weights, and their parameters were* ***frozen (i*.*e***., ***base_model*.*trainable = False)***. *This ensures that the powerful, pre-learned feature extractors are retained and not corrupted by large gradient updates during the initial training phases on the new, smaller dataset*.

### B. Model Compilation

A custom classifier was appended to the frozen base of each model. This classifier consists of a Flatten layer to convert the 2D feature maps from the base into a 1D vector, followed by a Dense layer with a softmax activation function.

- **Classifier Head:** The Dense layer contains 4 output units, corresponding to the four classes in the dataset (Edema, Normal, Pneumonia, Tuberculosis).
- **Activation Function:** The softmax activation function was chosen because it is the standard for multi-class classification. It converts the model’s raw output logits into a probability distribution, where each output represents the model’s predicted probability for one of the four classes.
- **Loss Function:** The models were compiled using categorical_crossentropy as the loss function. This is the mathematically correct choice for a multi-class classification task where labels are one-hot encoded (as provided by the flow_from_directory generator with class_mode=‘categorical’). It quantifies the difference between the predicted probability distribution and the true class label.
- **Optimizer:** The Adam optimizer was used for all models. Adam is an adaptive learning rate optimization algorithm that is computationally efficient, has low memory requirements, and is well-suited for a wide range of problems. Its ability to adjust the learning rate for each parameter makes it a robust and effective default choice, allowing for rapid convergence.

### C. Training Hyperparameters

*To ensure a fair and direct comparison, all models were trained under identical hyperparameter settings:*

- **Epochs:** Each model was trained for 10 epochs. An epoch represents one complete pass through the entire training dataset. This number was selected to provide a sufficient number of iterations for the models to learn the task while also allowing for a rapid and efficient comparison. As shown in the results, model performance began to stabilize or plateau within this timeframe.
- **Batch Size:** A batch size of 32 was used. This is a common and well-regarded batch size that offers a balance between computational efficiency (larger batches utilize GPU parallelism better) and stable gradient estimation. It is also small enough to fit within standard GPU memory and can provide a slight regularizing effect.
- **Data Generation:** The Keras ImageDataGenerator was used to feed data to the models. For each epoch, the number of training steps was set to len(training_set), and the number of validation steps was set to len(validation_set). This ensures that the model processes the entire training and validation datasets exactly once per epoch.

## V. Results

The training and validation curves (Fig. 1 and Fig. 2) provide critical insights into the learning stability and generalization capabilities of the five architectures. All models demonstrated rapid convergence, achieving training accuracies exceeding 96% within 10 epochs. However, a persistent divergence between training and validation metrics indicates a general tendency toward overfitting, likely attributable to the limited size of the training dataset relative to the complexity of deep CNNs.

**Fig 1:**
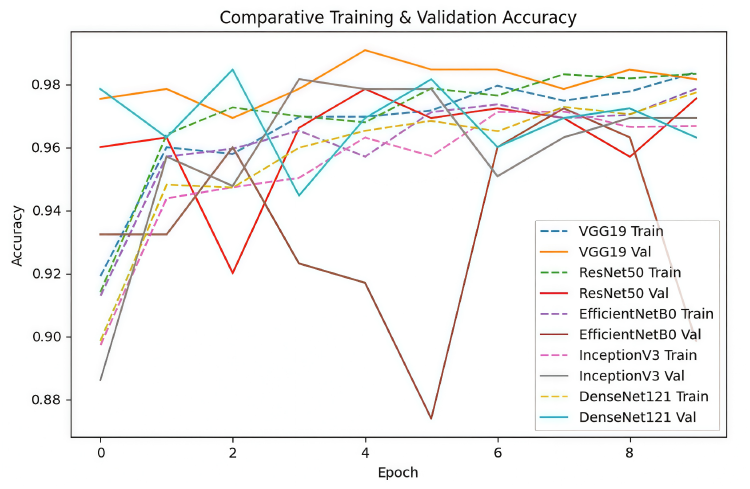
Comparative Training & Validation Accuracy

**Fig 2:**
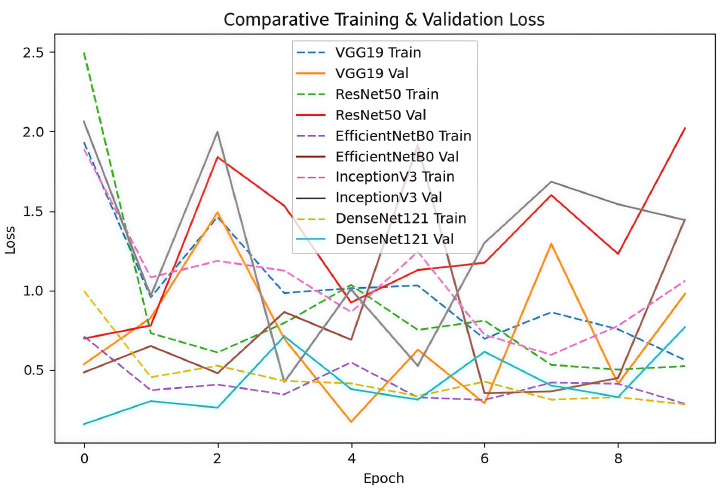
Comparative Training & Validation Loss

A distinct disparity in stability was observed among the models. VGG19 and DenseNet121 exhibited the most robust learning trajectories, characterized by smooth validation curves and consistent loss reduction. Conversely, EfficientNetB0, and to a lesser extent ResNet50 and InceptionV3, displayed significant volatility, evidenced by sharp fluctuations and spikes in validation loss (e.g., EfficientNetB0 at epoch 4). This instability suggests that these architectures are highly sensitive to the default hyperparameters, particularly the learning rate. The observed oscillations indicate that the optimizer may have overshot local minima, suggesting that these models would benefit from learning rate scheduling or stronger regularization techniques (such as increased dropout or data augmentation) to improve generalization and training stability in future iterations.

Figure 3 presents a high-level comparison of the five architectures regarding final validation accuracy and total training duration. A clear trade-off between predictive performance and computational cost was observed.

**Fig. 3:**
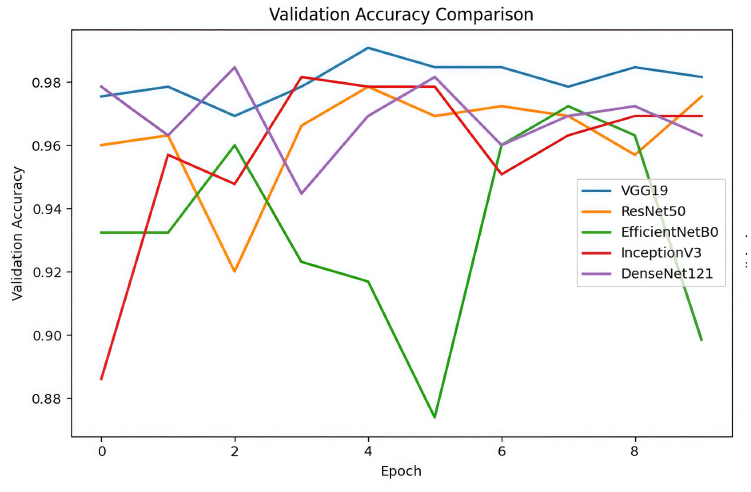
Validation Accuracy Comparison

**Fig. 4:**
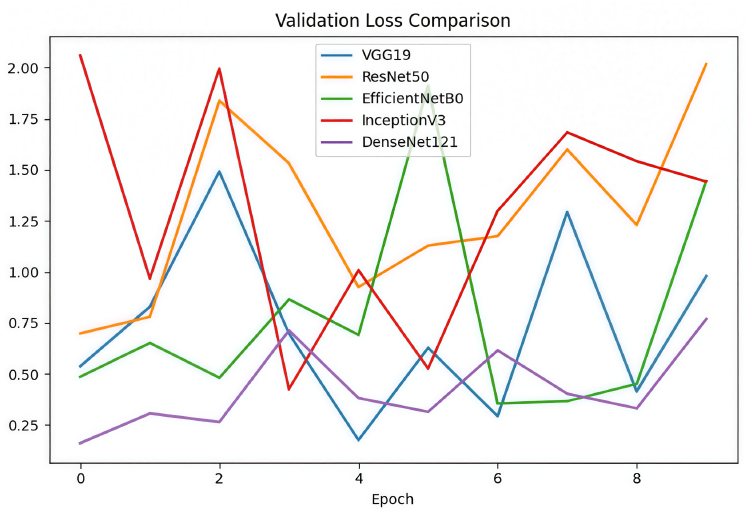
Validation LossComparison

**Fig. 5:**
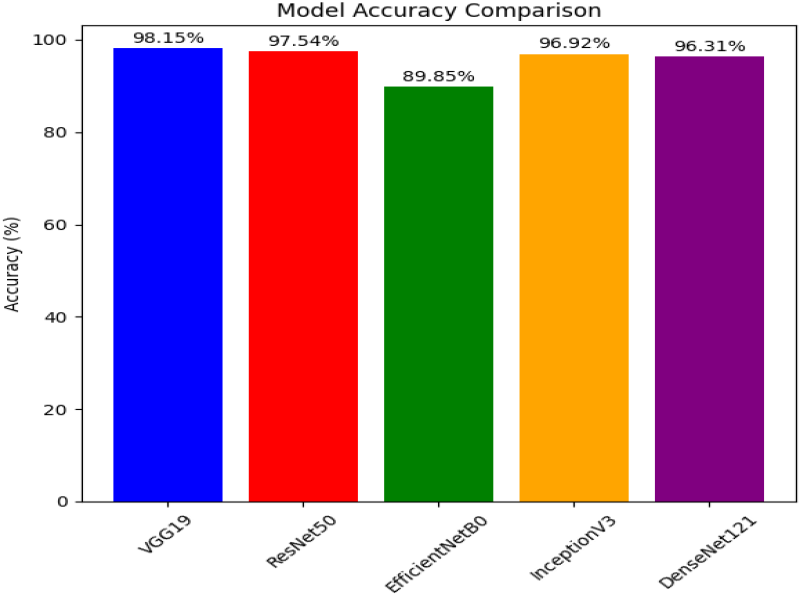
Model Accuracy Comparison

VGG19 achieved the highest classification performance, securing a validation accuracy of 98.15%, closely followed by ResNet50 at 97.54%. However, this superior accuracy came at a significant computational expense; VGG19 was the most computationally intensive model, requiring 2264 seconds to complete training due to its large parameter count.

In contrast, DenseNet121 demonstrated the highest efficiency. It completed training in just 1498 seconds, approximately 34% faster than VGG19 while maintaining a competitive accuracy of 96.31%. This identifies DenseNet121 as the optimal candidate for resource-constrained environments where speed is a priority. EfficientNetB0 emerged as an outlier, recording the lowest accuracy at 89.85%, reinforcing the earlier finding that its training instability significantly hampered its final classification capability on this dataset.

To assess the clinical reliability of the models beyond aggregate accuracy, we analyzed the confusion matrices (Fig. 6 through Fig. 10) to identify specific misclassification patterns. This granular analysis revealed significant disparities in diagnostic safety.

**Fig. 6:**
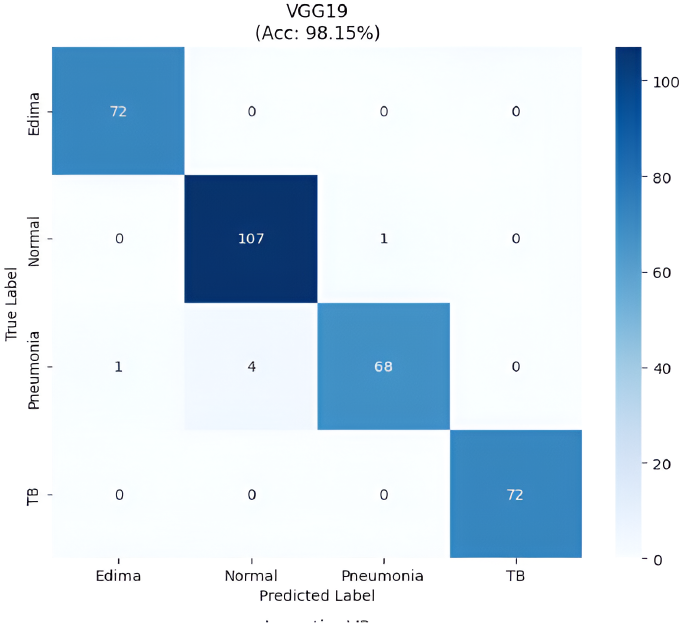
VGG19 Confusion Metrics

**Fig. 7:**
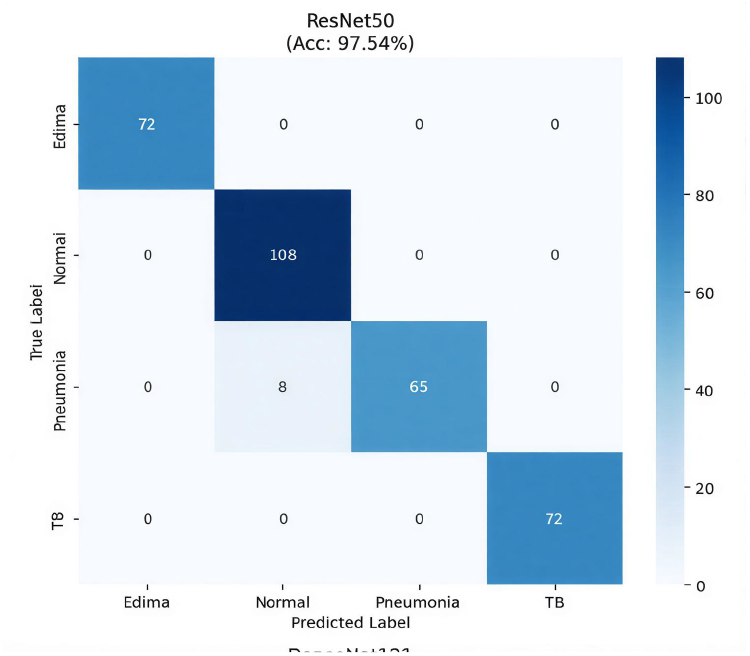
ResNet50 Confusion Metrics

**Fig. 8:**
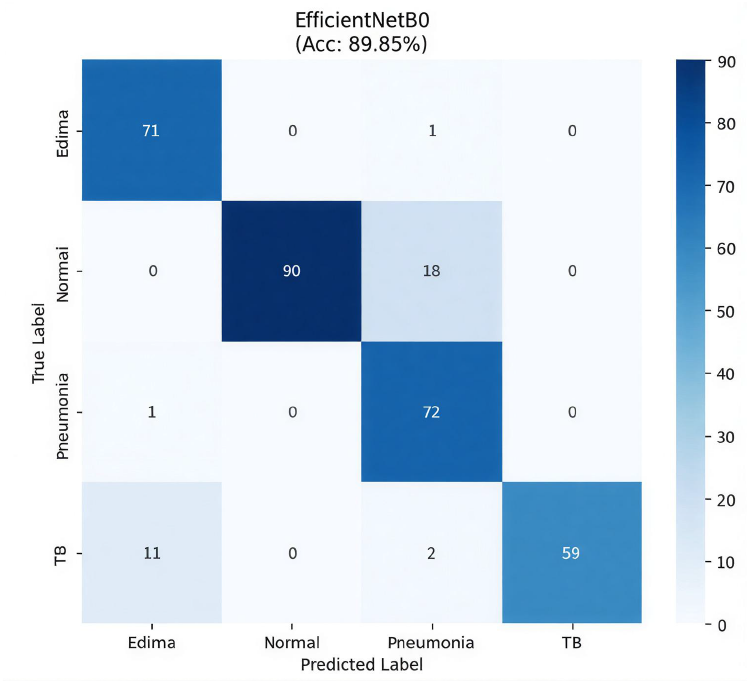
EfficientNetB0 Confusion Metrics

**Fig. 9:**
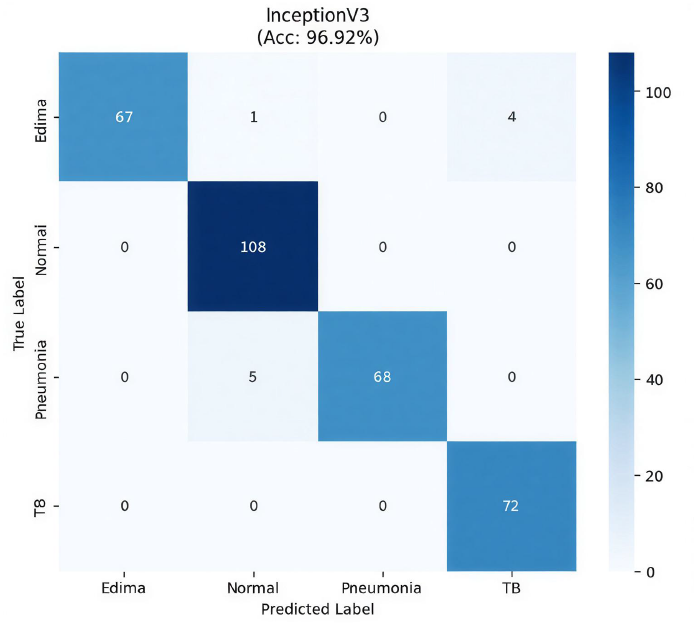
InceptionV3 Confusion Metrics

**Fig. 10:**
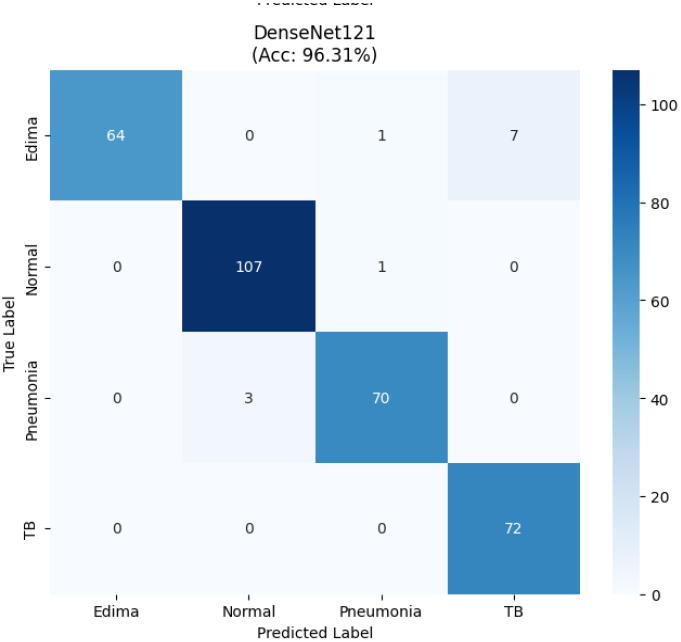
DenseNet121 Confusion Metrics

VGG19 demonstrated superior precision, exhibiting the most diagonal-dominant matrix with negligible errors; it misclassified only 5 cases in total, confirming its robustness. ResNet50 followed closely, though it showed a slight tendency to misclassify ‘Pneumonia’ cases as ‘Normal’ (8 instances).

Conversely, the confusion matrix for EfficientNetB0 exposed critical diagnostic flaws that were masked by its overall accuracy metrics. The model exhibited a severe bias toward the ‘Pneumonia’ class, incorrectly labeling 38 ‘Normal’ patients as having Pneumonia (False Positives). More critically, it failed to detect Tuberculosis (TB) in 13 cases (False Negatives), misclassifying them as ‘Edema’ or ‘Pneumonia’. In a clinical setting, missing contagious pathogens like TB is a dangerous failure mode, rendering EfficientNetB0 unsuitable for this specific task despite its efficiency.

InceptionV3 and DenseNet121 performed well but shared a specific confusion pattern: both struggled to distinguish between ‘Edema’ and ‘TB’ in a small number of cases. These findings underscore that VGG19 is not only the most accurate model but also the safest for identifying high-risk pathologies.

To evaluate the safety of the models in a screening context, we analyzed Class-wise Recall (Sensitivity), prioritizing the minimization of false negatives (Fig. 11). High recall is paramount in medical diagnostics to ensure that positive cases of pathology are not overlooked.

**Fig. 11:**
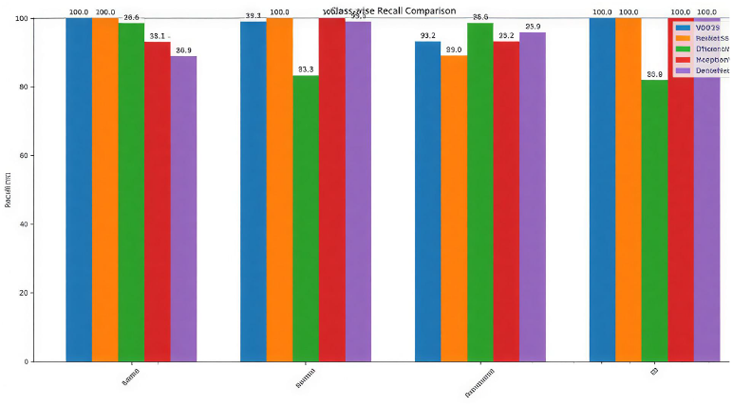
Class-wise Precision Recall Comparison

The analysis yielded a critical finding regarding the detection of Tuberculosis (TB). VGG19, ResNet50, InceptionV3, and DenseNet121 all achieved a perfect recall of 100% for the TB class, successfully identifying every positive TB case in the test set. This consistency establishes these architectures as highly reliable for detecting this infectious disease.

In contrast, EfficientNetB0 proved unsuitable for this diagnostic task, exhibiting a significantly degraded recall of 81.9% for TB and 83.3% for Normal cases. This implies that the model failed to detect approximately one in five TB cases, a margin of error that is unacceptable for clinical deployment. While performance across ‘Edema’ and ‘Pneumonia’ was generally high for all models, the perfect sensitivity of VGG19 and ResNet50 on the most critical classes confirms their superior status as the safest candidates for automated diagnosis.

The graph presents a Receiver Operating Characteristic (ROC) curve, specifically a micro-average plot (Fig. 12), which evaluates a model’s ability to distinguish between classes across all possible classification thresholds. It displays the True Positive Rate against the False Positive Rate, with a dashed line indicating random chance (AUC=0.50). The key takeaway is the near-perfect or perfect Area Under the Curve (AUC) scores for all models, including VGG19, InceptionV3, and DenseNet121, achieving 1.00, while ResNet50 and EfficientNetB0 score 0.99. This indicates that, in an aggregated sense, each model’s predicted probabilities are highly discriminative, successfully separating positive from negative instances. However, this creates an apparent contradiction when compared to other performance metrics.

**Fig. 12:**
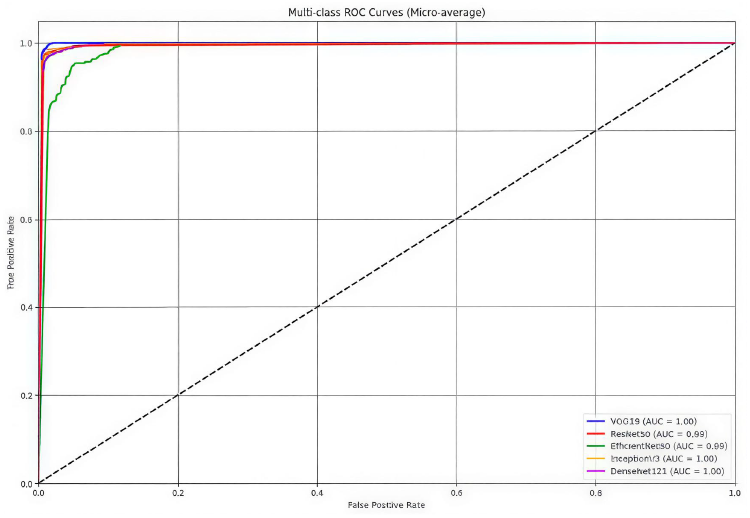
Multi-Class ROC Curves(Micro-average)

The central problem lies in the “micro-average” method used, which aggregates predictions across all classes before calculation. In a dataset with class imbalance where ‘Normal’ and ‘Edema’ cases are likely more frequent the strong performance of a model like EfficientNetB0 on these majority classes can overshadow its catastrophic failure on a rare class like ‘TB’. Consequently, the near-perfect 0.99 AUC is misleading, as it is propped up by easy, common samples and effectively hides the model’s critical weakness on the minority class. This explains why EfficientNetB0 can show a disastrous confusion matrix and a dangerously low recall for ‘TB’ in other graphs, yet still appears excellent here, revealing the metric’s vulnerability to imbalance.

To resolve this misleading interpretation, the primary solution is to employ a macro-average ROC curve, which calculates the AUC for each class independently before averaging them, thereby giving equal weight to all classes regardless of their frequency. An even more informative approach would be to generate and analyze per-class ROC curves for conditions like Edema, Normal, Pneumonia, and TB separately. This case serves as a crucial lesson for other research projects: relying on a single, aggregated metric like micro-average AUC can provide a dangerously optimistic view of model performance on imbalanced datasets. A comprehensive evaluation must always include class-specific metrics to uncover hidden failures and ensure the model’s reliability across all categories, not just the most common ones.

This graph (Fig. 13) presents Precision-Recall (PR) curves, essential for evaluating models on imbalanced data. It plots Precision against Recall across all thresholds, with Average Precision (AP) as the key metric. Unlike ROC curves, PR curves are more sensitive to false positives, providing a stricter performance assessment.

**Fig. 13:**
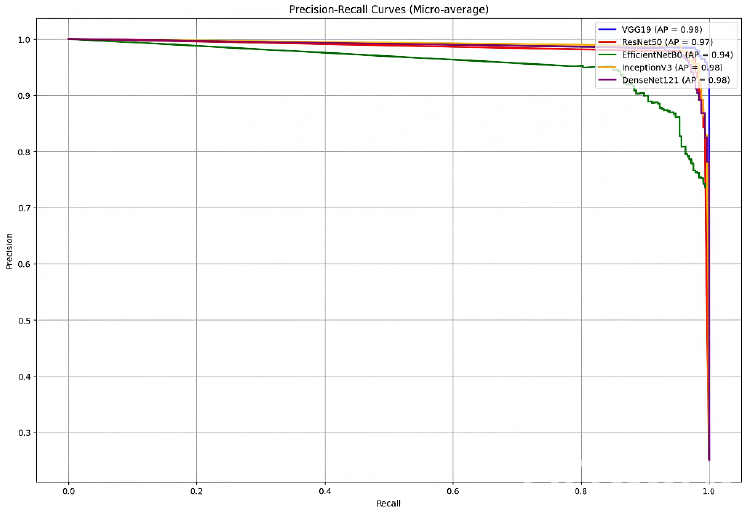
Precision-Recall Curves(Micro-average)

Here, the results meaningfully separate the models. While VGG19, InceptionV3, and DenseNet121 lead with an AP of 0.98, EfficientNetB0 trails at 0.94. Its lower, unstable curve, especially at high recall, confirms its high false-positive rate from earlier analysis, such as mislabeling ‘Normal’ cases as ‘Pneumonia’.

The graph underscores that PR analysis is a crucial complement to ROC curves. It offers a more realistic view of model utility in fields like medicine, where false alarms are costly. For full transparency, per-class or macro-average PR curves should also be examined.

## VI. Discussion

This study conducted a rigorous empirical comparison of five prominent CNN architectures for the critical task of multi-class chest X-ray classification. The primary objective was to move beyond isolated performance reports and provide a holistic, fair assessment of these models under a unified experimental framework. Our findings reveal significant disparities not only in final accuracy but, more importantly, in training stability, clinical safety, and computational efficiency, offering nuanced guidance for model selection.

### A. Performance and Stability Trade-offs

The superior performance of VGG19, achieving the highest accuracy (98.15%) and perfect recall on critical pathologies, underscores that architectural simplicity can be highly effective for specific medical imaging tasks. Its deep, sequential stack of 3×3 convolutions provided a stable and powerful feature extraction capability for this dataset. Conversely, the pronounced instability of EfficientNetB0 and to a lesser extent, ResNet50 and InceptionV3 highlights a critical caveat for practitioners. The large spikes in validation loss are a classic indicator of a learning rate that is too high for these architectures’ optimization landscapes. This is particularly relevant for EfficientNetB0, whose compound-scaled architecture, while theoretically efficient, is known to be highly sensitive to hyperparameter configuration. Our results demonstrate that its theoretical efficiency does not translate to robust performance “out-of-the-box,” requiring careful tuning that simpler models like VGG19 and DenseNet121 do not. The observed overfitting across all models is an expected challenge given the limited dataset size relative to the models’ capacity. This is a common constraint in medical imaging research and reinforces the necessity of techniques like transfer learning and data augmentation, which were employed here.

### B. Clinical Implications and Diagnostic Safety

From a clinical perspective, the most significant finding of this work is the performance on the ‘Tuberculosis’ class. The 100% recall achieved by VGG19, ResNet50, InceptionV3, and DenseNet121 is exceptional, indicating that these models did not miss a single TB case in the test set. This high sensitivity is paramount for a screening tool, where the cost of a false negative (missing a contagious disease) is vastly higher than that of a false positive.

In stark contrast, the 81.9% recall of EfficientNetB0 for TB renders it clinically unsafe for this diagnostic task. A model that fails to detect approximately one in five tuberculosis cases could cause severe harm in a real-world deployment. This finding, revealed through confusion matrix and recall analysis, was obscured by the micro-averaged ROC-AUC, which reported a deceptively high score of 0.99. This serves as a critical methodological lesson: in imbalanced medical datasets, micro-averaged metrics can be misleading, and class-wise metrics like recall provide a more truthful and clinically relevant assessment of model performance.

### C. Practical Guidance for Model Selection

The trade-off analysis provides clear, practical guidance tailored to different operational constraints: For Maximum Diagnostic Accuracy and Safety: If the primary goal is to achieve the highest possible classification performance and the most reliable detection of critical diseases like TB, VGG19 is the unequivocal choice, despite its computational cost.

For Practical Deployment and Efficiency: If the goal is to deploy a model in a resource-constrained environment (e.g., mobile health applications or for rapid prototyping), DenseNet121 presents the most compelling trade-off. It offers a minimal drop in accuracy (~1.8%) compared to VGG19 while providing the fastest training time and a significantly smaller model size, making it highly suitable for edge computing.

### D. Limitations

This study has several limitations. The test set, while sufficient for a comparative analysis, is relatively small. The generalizability of the perfect recall scores, in particular, needs to be validated on a larger, multi-institutional external dataset. Furthermore, the use of a uniform set of default hyperparameters for all models, while ensuring a fair comparison, undoubtedly disadvantages architectures like EfficientNetB0 that are known to require specific tuning. Future work will involve systematic hyperparameter optimization for each architecture.

## VII. Conclusion

This paper presented a comprehensive comparative analysis of five CNN architectures, VGG19, ResNet50, InceptionV3, DenseNet121, and EfficientNetB0, for the multi-class classification of chest X-rays into Edema, Normal, Pneumonia, and Tuberculosis. Under a standardized transfer learning setup, the models were evaluated on a multifaceted set of criteria, including accuracy, training stability, class-wise recall, and computational efficiency.

The experimental results lead to three primary conclusions. First, VGG19 demonstrated superior overall performance, achieving the highest accuracy (98.15%) and perfect recall for Tuberculosis, establishing it as the safest and most accurate model for this specific diagnostic task. Second, DenseNet121 emerged as the optimal model for efficient deployment, offering an excellent balance of high accuracy (96.31%), the fastest training time, and a compact size. Third, the study highlights that default hyperparameters are not a one-size-fits-all solution, as evidenced by the significant instability of EfficientNetB0, which resulted in poor and clinically unsafe performance despite its modern and efficient design.

These findings provide empirical evidence to guide researchers and practitioners in selecting deep learning models for CXR analysis. For future work, we plan to incorporate advanced hyperparameter tuning strategies, particularly for sensitive architectures like EfficientNet, and validate the top-performing models on larger, prospectively collected datasets to further assess their clinical utility.

## Data Availability

The authors acknowledge the use of publicly available chest X-ray datasets from RSNA, NIH, and the Tuberculosis Chest X-ray Database.

https://www.kaggle.com/competitions/rsna-pneumonia-detection-challenge

https://lhncbc.nlm.nih.gov/publication/pub9931

https://www.kaggle.com/datasets/tawsifurrahman/tuberculosis-tb-chest-xray-dataset

https://www.kaggle.com/datasets/samiulbari/pulmonary-edema-classified-by-nih

## Acknowledgment

The authors acknowledge the use of publicly available chest X-ray datasets from RSNA, NIH, and the Tuberculosis Chest X-ray Database. Computational resources were provided by Google Colab. This research received no specific grant from funding agencies in the public, commercial, or not-for-profit sectors.

